# Stroke Volume Reserve Is an Independent Predictor of Survival and Need for Advanced Therapies in Systolic Heart Failure Patients

**DOI:** 10.1101/2019.12.21.19015610

**Authors:** Ibrahim Selevany, Florin Vaida, Eric D. Adler, Paul J. Kim, Timothy Morris

## Abstract

**Background:** Current predictors of clinical deterioration in patients with heart-failure with reduced ejection fraction (HFrEF) have limited accuracy. We investigated whether stroke volume reserve index (SVRI) would predict 1-year mortality and need for advanced therapies in HFrEF patients.

**Methods:** We retrospectively studied a consecutive series of 104 ambulatory HFrEF patients (59±11 years, 89% men) at our center who underwent cardiopulmonary exercise testing (CPET) from January 2017 to September 2018. The primary outcome was defined as heart-transplantation, urgent left ventricular assist device (LVAD) implantation or death within 1 year of evaluation. SVRI was estimated at rest and at anaerobic threshold (AT) by VO_2_ (oxygen consumption)/pulse and previously validated estimates of C[a-v]O_2_ (arteriovenous difference in oxygen content). Multivariate regression selected the optimal predictors from clinical, CPET and right heart catheterization data.

**Results:** Among HFrEF patients studied, 39 (37.5%) deteriorated: 16 required heart-transplantation, 8 required LVADs and 15 died. SVRI 128% or less was the best predictor (OR 6.69, p=0.001), followed by an invasively determined resting cardiac index 2 L/min/m^2^ or less (OR 3.18, p=0.02), and peak VO_2_ below the International Society for Heart and Lung Transplantation (ISHLT) cut-off (OR 4.15, p=0.01). A scoring system derived from the SVRI (less than 128%: 2 points), peak VO_2_ (below the ISHLT cut-off: 1 point), and cardiac index (less than 2 L/min/m^2^: 1 point) predicted clinical deterioration with a sensitivity of 64% and specificity of 81%.

**Conclusions:** SVRI is a non-invasive measurement that may predict deterioration in HFrEF patients more accurately than currently recommended predictors.

## INTRODUCTION

The morbidity and mortality of patients with heart-failure have improved considerably over recent years.^1-3^ However, there remain patients who continue to progress in their heart-failure course and require evaluation for advanced therapies.^4^ These patients are considered to be in advanced or American College of Cardiology/American Heart Association Stage D heart-failure.^5^ Exercise tolerance, typically represented as oxygen consumption (VO_2_) less than 12-14 ml/kg/min, is a commonly recommended indicator for advanced therapies.^6, 7^ However, peak VO_2_ has only a limited accuracy to predict serious clinical deterioration in patients with heart failure.^8^ Other exercise parameters such as VE/VCO_2_ slope, VO_2_ at AT and peak VO_2_ pulse also have limited accuracy of predicting deterioration in heart failure patients.^8-15^

We recently reported that the ratio of the stroke volume (SV) at AT to the SV at rest (stroke volume reserve index, or SVRI) measured during cardiopulmonary exercise testing (CPET) reflects right ventricular dysfunction after acute pulmonary embolism.^16^ The SVRI is measured non-invasively using a method developed by Stringer et al.^17^ and matches invasively-measured values in normal subjects and heart failure patients.^16, 18-21^ Because the cardiac response to exercise is an important determinant of outcome in patients with cardiomyopathy^22, 23^, and because SVRI reflects SV at two standardized points during exercise, we reasoned that decreased SVRI would predict clinical deterioration more accurately than currently used CPET parameters.^7, 24-26^ To test our hypothesis, we analyzed the records of a retrospective series of HFrEF patients who had performed CPET, to determine the accuracy with which decreased SVRI predicted death, the need for heart-transplantation or urgent LVAD implantation.

## METHODS

### Study Population and Design

We retrospectively analyzed all HFrEF patients who underwent CPET at the University of California, San Diego (UC San Diego) physiology laboratory for advanced therapies evaluation between January 1, 2017 and September 30, 2018. We defined HFrEF as left ventricular ejection fraction (LVEF) of 35% or less by echocardiography. Patients less than 18 years old, with congenital heart disease, planned coronary revascularization, previous transplantation, intravenous inotropic therapy and those who did not reach AT during CPET were excluded. The UC San Diego Human Research Protections Program approved the retrospective analysis of data without the need for informed consent. All of the authors have full access to the data in the study and take full responsibility for its integrity and the analysis.

### Exercise Protocol

Incremental symptom-limited CPET was performed using a V-max metabolic cart (CareFusion, San Diego, CA), as previously described.^16^ All patients performed exercise testing on a treadmill. The treadmill protocols had been clinically validated within each weight interval to produce a uniform increase in mechanical work and metabolic energy expenditure per incremental step. Exhaled gases, electrocardiography and pulse oximetry were measured continuously. AT was determined by a board certified pulmonologist (TM) with the V-slope method.^27^

### SV at rest and at AT

We estimated SVRI as previously described.^16^ SV during rest and AT were estimated with the equation

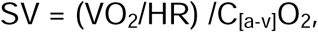

where SV = stroke volume, VO_2_ = oxygen consumption, HR = heart rate (both measured simultaneously), (O_2_·pulse = VO_2_/HR) and C[a-v]O_2_ = arteriovenous oxygen content difference.^17^ C_[a-v]_O_2_, when previously measured by arterial and pulmonary artery blood sampling in healthy subjects and in patients at various stages of heart failure, is approximately 6.1 ml O_2_ / 100 ml blood at rest and 11.3 ml O_2_ / 100 ml blood at AT.^17-21^ The ratio between the two C_[a-v]_O_2_ values also corresponds to the ratios we observed in patients with pulmonary hypertension who underwent exercise testing in our center’s catheterization laboratory with pulmonary artery and arterial catheters in place.^16^ Based on those observations, we estimated the SVRI as:

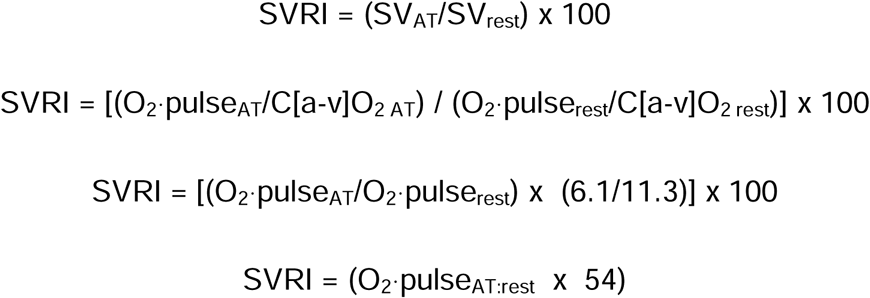

where O_2_·pulse_AT:rest_ = the ratio of the O_2_·pulse at AT to the O_2_·pulse at rest.^17^ The SVRI measured with this method is approximately 140% +/- 12%^17-19, 21^ among healthy subjects, which corresponds to an O_2_·pulse_AT:rest_ of 2.59 +/- 0.22.^17, 28^ Before data collection, we pre-defined an abnormally low SVRI to be 128% or lower based on the variability observed in in healthy subjects who underwent invasive studies.^16, 17^

### Clinical Characteristics

From the clinical data collected by review of the electronic medical records prior to the observation period, we calculated three clinically validated risk stratification scores, namely the Heart Failure Survival Score (HFSS)^29^, the Seattle Heart Failure Model (SHFM) score^30^, and the Kato model score.^31^

### Right heart catheterization

We also included data from all right heart catheterization procedures that had been performed for clinical reasons within 30 days of the CPET. Resting cardiac output (CO) and cardiac index was determined by the Fick method. Pulmonary vascular resistance (PVR) was calculated as: PVR = [mean pulmonary artery pressure (MPA) - pulmonary capillary wedge pressure / CO. The right ventricular stroke work index (RVSWI) was calculated as (MPA - mean RAP, right atrial pressure) × SV index × 0.0136 (g-m2/beat).

### Outcomes and Data Acquisition

The primary outcome was a composite of death, the need for either urgent LVAD implantation or heart-transplantation one year following CPET. A trained investigator (IS) determined the outcomes by a structured chart review, as well as structured telephone interviews for patients that were not seen in the hospital system within the past three months. At the time of each patient’s evaluation by the advanced therapies selection committee, the committee was not informed of the SVRI and thus it did not influence the committee’s decisions regarding the patient’s candidacy for advanced therapies.

### Statistical Analysis

The primary analysis was to determine the accuracy of SVRI for predicting the 1-year composite outcome, compared to other candidate predictors. The predictors were identified using single- and multi-predictor logistic regression. Multi-predictor logistic regression used backwards model selection, starting with the covariates significant at p<0.05 in single-predictor models, and with a p<0.05 threshold for inclusion.

As an additional step, the predictors from the final multi-predictor model were dichotomized using pre-defined cut-offs. The cut-off value for SVRI was pre-determined to be 128% or lower as previously described. The cut-off value for invasively determined resting cardiac index was chosen to minimize the Euclidian distance from the receiver-operating-characteristic (ROC) curve to the upper-left corner of the ROC graph, in effect maximizing a combination of sensitivity and specificity. The cut-off values for peak VO_2_ (12 or 14 ml/kg/min in the presence of a β-blocker) followed the 2016 ISHLT guidelines for heart-transplant listing.^7^

We also compared the demographic and clinical characteristics of the two cohorts with SVRI above and below the cut-off. Comparison of variables used independent Student’s t-test for continuous variables, and Fisher’s exact test for binary variables.

Based on the final multi-predictor logistic regression model, which included the dichotomized SVRI, peak VO_2_, and resting cardiac index, we created a “SVRI risk score”, which uses simple integer weights for each of the three covariates, based on their model coefficient (i.e., log odds ratio), with the lowest coefficient receiving 1 point, as a reference value. The reason for using the logistic regression model coefficients as a basis for the score is that in the predictive model the risk score is linearly associated with the log-odds of the event. This method is similar but not identical to that of Kato et al.^31^

The time-to-event distributions were generated separately for the two groups based on SVRI below and above the threshold, and for the four groups based on the cross-classification of SVRI below/above threshold and peak VO_2_ below/above threshold, using the Kaplan-Meier estimator. The relative risk in the different groups was assessed via the hazard ratio from the Cox proportional hazards model, and group comparison was based on the log-rank test.

In all analyses p < 0.05 was considered significant; two-sided hypothesis tests and confidence intervals were used throughout. Statistical analyses were performed using Prism 8 for Windows (GraphPad Software, Inc, San Diego, CA) and Excel’s XLMiner Analysis ToolPak (Microsoft Excel; Microsoft Corp, Bellevue, WA).

## RESULTS

### Clinical Characteristics and 1-year Outcomes

Between January 1, 2017 and September 30, 2018, 129 patients with LVEF ≤ 35% by echocardiography underwent CPET at our center. 25 patients were excluded: nine had congenital heart disease, six were scheduled for coronary revascularization, three had previous heart transplantation, two were being treated with intravenous inotropic therapy and five did not reach AT during CPET. (Three of the five patients who failed to reach AT died within one year: a phenomenon also observed by Agostoni and colleagues.^32^) The remaining 104 patients constituted our study group.

The clinical characteristics of the study population are summarized in Table 1. The mean age of the patients was 59 years, 89% were male, and 51% had an ischemic etiology. 88% of patients were on **β**-blocker therapy, 25% also requiring digoxin or ivabradine, 95% on either ACEI/ARB, ARNI or isosorbide dinitrate/hydralazine therapy, and 84% received either an ICD or CRT. All patients had symptomatic heart failure, with a mean NYHA class of 2.8 ± 0.6. Mean peak VO_2_ at the time of heart-transplant evaluation was 15.5 ml/kg/min, mean LVEF was 22.3% and mean SVRI was 1.2. Right heart catheterization had been performed for 82 of the 104 patients. The mean RAP was 8.9 mmHg, PCWP was 18.2 mmHg, PVR 2.8 Wood units and resting cardiac index of 2.2 liters/min/m^2^.

**Table 1.**
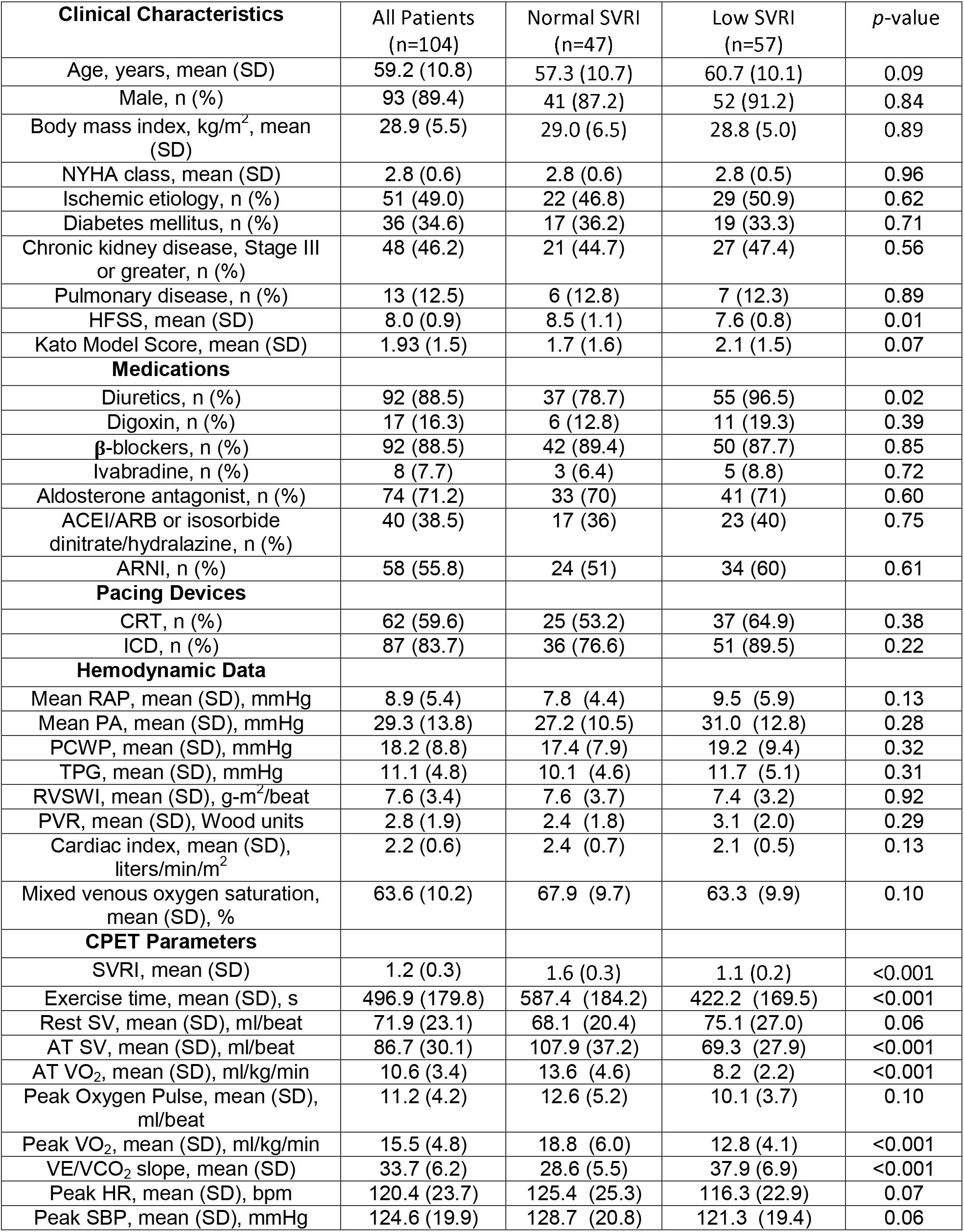
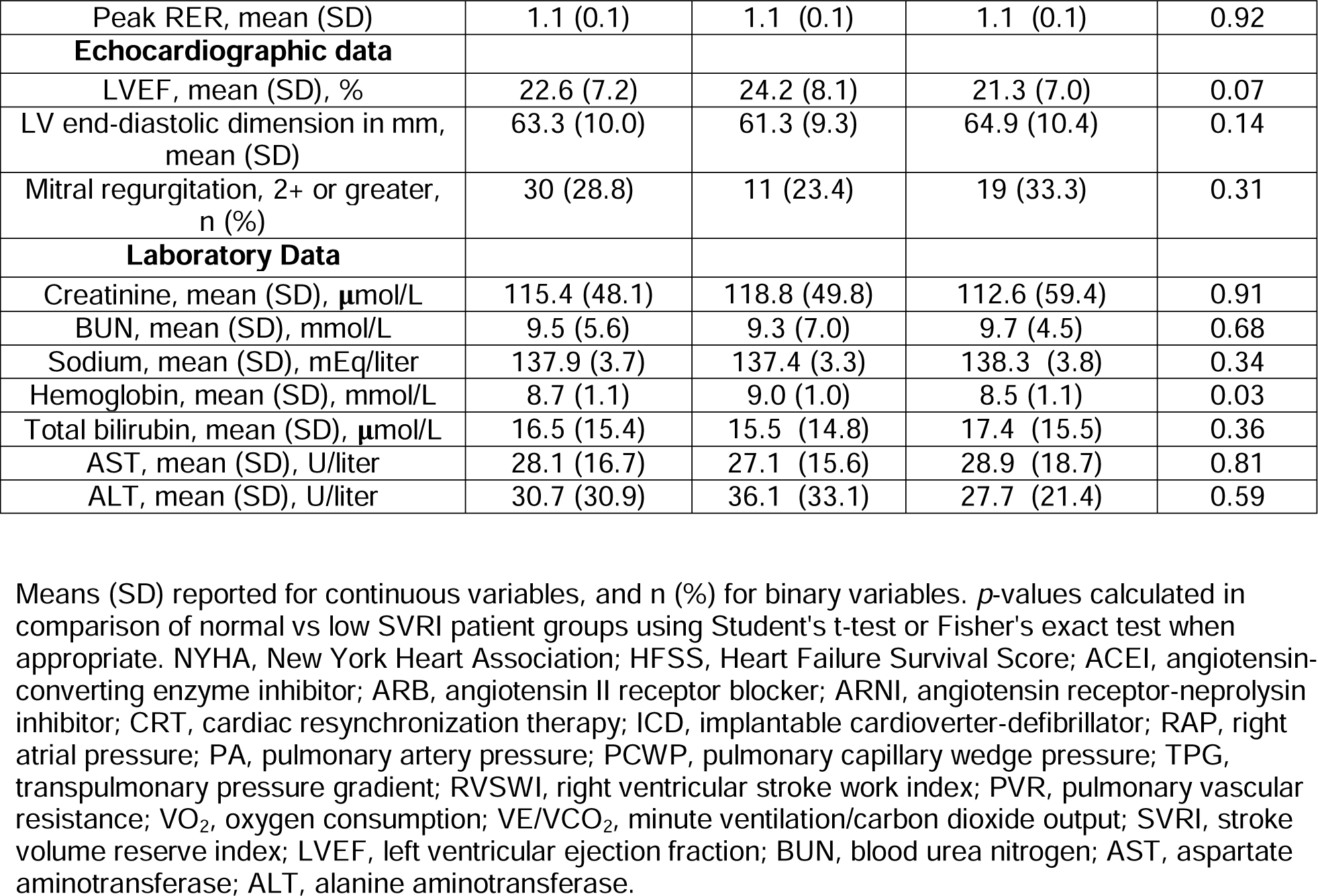
Clinical Characteristics of Patients at Time of CPET

| **Clinical Characteristics** | All Patients<br>(n=104) | Normal SVRI<br>(n=47) | Low SVRI<br>(n=57) | *p*-value |
| --- | --- | --- | --- | --- |
| Age, years, mean (SD) | 59.2 (10.8) | 57.3 (10.7) | 60.7 (10.1) | 0.09 |
| Male, n (%) | 93 (89.4) | 41 (87.2) | 52 (91.2) | 0.84 |
| Body mass index, kg/m<sup>2</sup>, mean (SD) | 28.9 (5.5) | 29.0 (6.5) | 28.8 (5.0) | 0.89 |
| NYHA class, mean (SD) | 2.8 (0.6) | 2.8 (0.6) | 2.8 (0.5) | 0.96 |
| Ischemic etiology, n (%) | 51 (49.0) | 22 (46.8) | 29 (50.9) | 0.62 |
| Diabetes mellitus, n (%) | 36 (34.6) | 17 (36.2) | 19 (33.3) | 0.71 |
| Chronic kidney disease, Stage III or greater, n (%) | 48 (46.2) | 21 (44.7) | 27 (47.4) | 0.56 |
| Pulmonary disease, n (%) | 13 (12.5) | 6 (12.8) | 7 (12.3) | 0.89 |
| HFSS, mean (SD) | 8.0 (0.9) | 8.5 (1.1) | 7.6 (0.8) | 0.01 |
| Kato Model Score, mean (SD) | 1.93 (1.5) | 1.7 (1.6) | 2.1 (1.5) | 0.07 |
| **Medications** |  |  |  |  |
| Diuretics, n (%) | 92 (88.5) | 37 (78.7) | 55 (96.5) | 0.02 |
| Digoxin, n (%) | 17 (16.3) | 6 (12.8) | 11 (19.3) | 0.39 |
| β-blockers, n (%) | 92 (88.5) | 42 (89.4) | 50 (87.7) | 0.85 |
| Ivabradine, n (%) | 8 (7.7) | 3 (6.4) | 5 (8.8) | 0.72 |
| Aldosterone antagonist, n (%) | 74 (71.2) | 33 (70) | 41 (71) | 0.60 |
| ACEI/ARB or isosorbide dinitrate/hydralazine, n (%) | 40 (38.5) | 17 (36) | 23 (40) | 0.75 |
| ARNI, n (%) | 58 (55.8) | 24 (51) | 34 (60) | 0.61 |
| **Pacing Devices** |  |  |  |  |
| CRT, n (%) | 62 (59.6) | 25 (53.2) | 37 (64.9) | 0.38 |
| ICD, n (%) | 87 (83.7) | 36 (76.6) | 51 (89.5) | 0.22 |
| **Hemodynamic Data** |  |  |  |  |
| Mean RAP, mean (SD), mmHg | 8.9 (5.4) | 7.8 (4.4) | 9.5 (5.9) | 0.13 |
| Mean PA, mean (SD), mmHg | 29.3 (13.8) | 27.2 (10.5) | 31.0 (12.8) | 0.28 |
| PCWP, mean (SD), mmHg | 18.2 (8.8) | 17.4 (7.9) | 19.2 (9.4) | 0.32 |
| TPG, mean (SD), mmHg | 11.1 (4.8) | 10.1 (4.6) | 11.7 (5.1) | 0.31 |
| RVSWI, mean (SD), g·m<sup>2</sup>/beat | 7.6 (3.4) | 7.6 (3.7) | 7.4 (3.2) | 0.92 |
| PVR, mean (SD), Wood units | 2.8 (1.9) | 2.4 (1.8) | 3.1 (2.0) | 0.29 |
| Cardiac index, mean (SD), liters/min/m<sup>2</sup> | 2.2 (0.6) | 2.4 (0.7) | 2.1 (0.5) | 0.13 |
| Mixed venous oxygen saturation, mean (SD), % | 63.6 (10.2) | 67.9 (9.7) | 63.3 (9.9) | 0.10 |
| **CPET Parameters** |  |  |  |  |
| SVRI, mean (SD) | 1.2 (0.3) | 1.6 (0.3) | 1.1 (0.2) | <0.001 |
| Exercise time, mean (SD), s | 496.9 (179.8) | 587.4 (184.2) | 422.2 (169.5) | <0.001 |
| Rest SV, mean (SD), ml/beat | 71.9 (23.1) | 68.1 (20.4) | 75.1 (27.0) | 0.06 |
| AT SV, mean (SD), ml/beat | 86.7 (30.1) | 107.9 (37.2) | 69.3 (27.9) | <0.001 |
| AT VO<sub>2</sub>, mean (SD), ml/kg/min | 10.6 (3.4) | 13.6 (4.6) | 8.2 (2.2) | <0.001 |
| Peak Oxygen Pulse, mean (SD), ml/beat | 11.2 (4.2) | 12.6 (5.2) | 10.1 (3.7) | 0.10 |
| Peak VO<sub>2</sub>, mean (SD), ml/kg/min | 15.5 (4.8) | 18.8 (6.0) | 12.8 (4.1) | <0.001 |
| VE/VCO<sub>2</sub> slope, mean (SD) | 33.7 (6.2) | 28.6 (5.5) | 37.9 (6.9) | <0.001 |
| Peak HR, mean (SD), bpm | 120.4 (23.7) | 125.4 (25.3) | 116.3 (22.9) | 0.07 |
| Peak SBP, mean (SD), mmHg | 124.6 (19.9) | 128.7 (20.8) | 121.3 (19.4) | 0.06 |
| Peak RER, mean (SD) | 1.1 (0.1) | 1.1 (0.1) | 1.1 (0.1) | 0.92 |
| **Echocardiographic data** |  |  |  |  |
| LVEF, mean (SD), % | 22.6 (7.2) | 24.2 (8.1) | 21.3 (7.0) | 0.07 |
| LV end-diastolic dimension in mm, mean (SD) | 63.3 (10.0) | 61.3 (9.3) | 64.9 (10.4) | 0.14 |
| Mitral regurgitation, 2+ or greater, n (%) | 30 (28.8) | 11 (23.4) | 19 (33.3) | 0.31 |
| **Laboratory Data** |  |  |  |  |
| Creatinine, mean (SD), μmol/L | 115.4 (48.1) | 118.8 (49.8) | 112.6 (59.4) | 0.91 |
| BUN, mean (SD), mmol/L | 9.5 (5.6) | 9.3 (7.0) | 9.7 (4.5) | 0.68 |
| Sodium, mean (SD), mEq/liter | 137.9 (3.7) | 137.4 (3.3) | 138.3 (3.8) | 0.34 |
| Hemoglobin, mean (SD), mmol/L | 8.7 (1.1) | 9.0 (1.0) | 8.5 (1.1) | 0.03 |
| Total bilirubin, mean (SD), μmol/L | 16.5 (15.4) | 15.5 (14.8) | 17.4 (15.5) | 0.36 |
| AST, mean (SD), U/liter | 28.1 (16.7) | 27.1 (15.6) | 28.9 (18.7) | 0.81 |
| ALT, mean (SD), U/liter | 30.7 (30.9) | 36.1 (33.1) | 27.7 (21.4) | 0.59 |

Means (SD) reported for continuous variables, and n (%) for binary variables. *p*-values calculated in comparison of normal vs low SVRI patient groups using Student's t-test or Fisher's exact test when appropriate. NYHA, New York Heart Association; HFSS, Heart Failure Survival Score; ACEI, angiotensin-converting enzyme inhibitor; ARB, angiotensin II receptor blocker; ARNI, angiotensin receptor-neprilysin inhibitor; CRT, cardiac resynchronization therapy; ICD, implantable cardioverter-defibrillator; RAP, right atrial pressure; PA, pulmonary artery pressure; PCWP, pulmonary capillary wedge pressure; TPG, transpulmonary pressure gradient; RVSWI, right ventricular stroke work index; PVR, pulmonary vascular resistance; $VO_2$ , oxygen consumption; VE/VCO<sub>2</sub>, minute ventilation/carbon dioxide output; SVRI, stroke volume reserve index; LVEF, left ventricular ejection fraction; BUN, blood urea nitrogen; AST, aspartate aminotransferase; ALT, alanine aminotransferase.

Among the 104 patients included in the study, 39 (37.5%) reached the primary composite end-point in the year subsequent to the index CPET: 16 patients (15.4%) required heart transplantation, 8 (7.7%) required urgent LVAD implantation and 15 (14.4%) died within 1 year.

### Comparison of Patients Dichotomized by SVRI

Patients who were above and below the predetermined cut-off of SVRI≤128% were similar in clinical characteristics, except that the low SVRI group had slightly lower HFSS (7.6 +/- 0.8 vs 8.5 +/- 1.1, p = 0.01), more diuretic use (55% vs 37%, p = 0.02) and slightly lower hemoglobin level (8.5 mmol/L vs 9.0 mmol/L, p = 0.03) (Table 1). Although there were no significant differences in right heart catheterization values, CPETs in the low SVRI group had lower exercise times (422.2 sec vs 587.4 sec, p < 0.001), lower VO_2_ at AT (AT VO_2_; 8.2 ml/kg/min vs 13.6 ml/kg/min, p < 0.001), lower peak VO_2_, (12.8 ml/kg/min vs 18.8 ml/kg/min, p < 0.001) and higher VE/VCO_2_ slopes (37.9 vs 28.6, p < 0.001).

### SVRI as a Predictor of Adverse Outcome

The 1-year event-free survival rate for patients with SVRI 128% or less was significantly lower than for those with higher SVRI (44% vs 85%, p < 0.001) (Fig. 1). As illustrated in Figure 2, the C-statistic for SVRI (0.865, 95% CI 0.795, 0.936) was greater than the C-statistic for peak VO_2_ (0.774, 95% CI 0.684, 0.865), AT VO_2_ (0.676, 95% CI 0.571, 0.781) and PCWP (0.678, 95% CI 0.573, 0.783).

**Figure 1.**
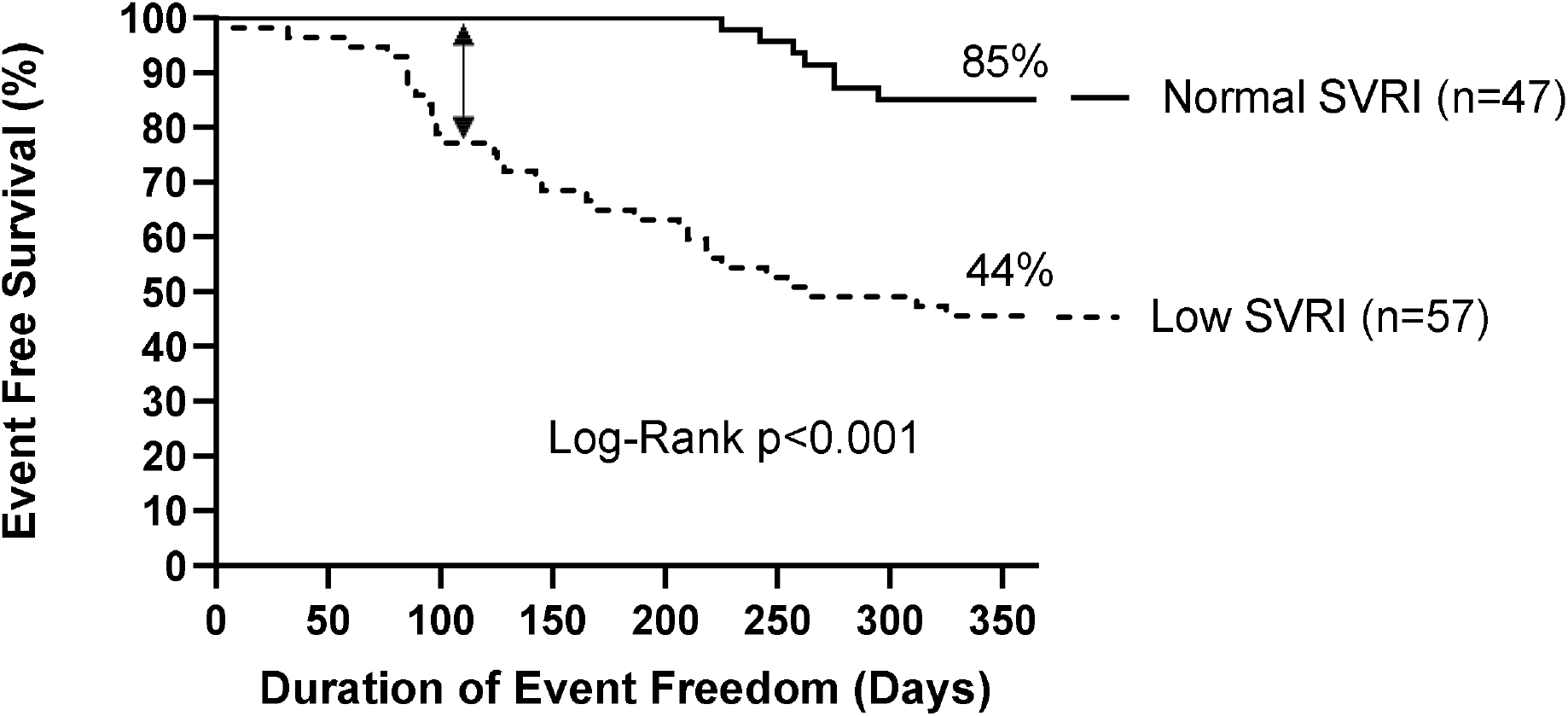
Kaplan-Meier curves of heart-failure with reduced ejection fraction patients after CPET classified as low SVRI (SVRI≤128%) and normal SVRI (SVRI>128%). The primary composite end-point was set as death, urgent left ventricular assist device implantation, or heart-transplantation. Patients with normal SVRI showed significantly higher 1-year event free survival compared to low SVRI patients. This difference in event free survival is seen as early as 100 days after the CPET (double sided arrow). CPET, cardiopulmonary exercise testing; SVRI, stroke volume reserve index.

**Figure 2.**
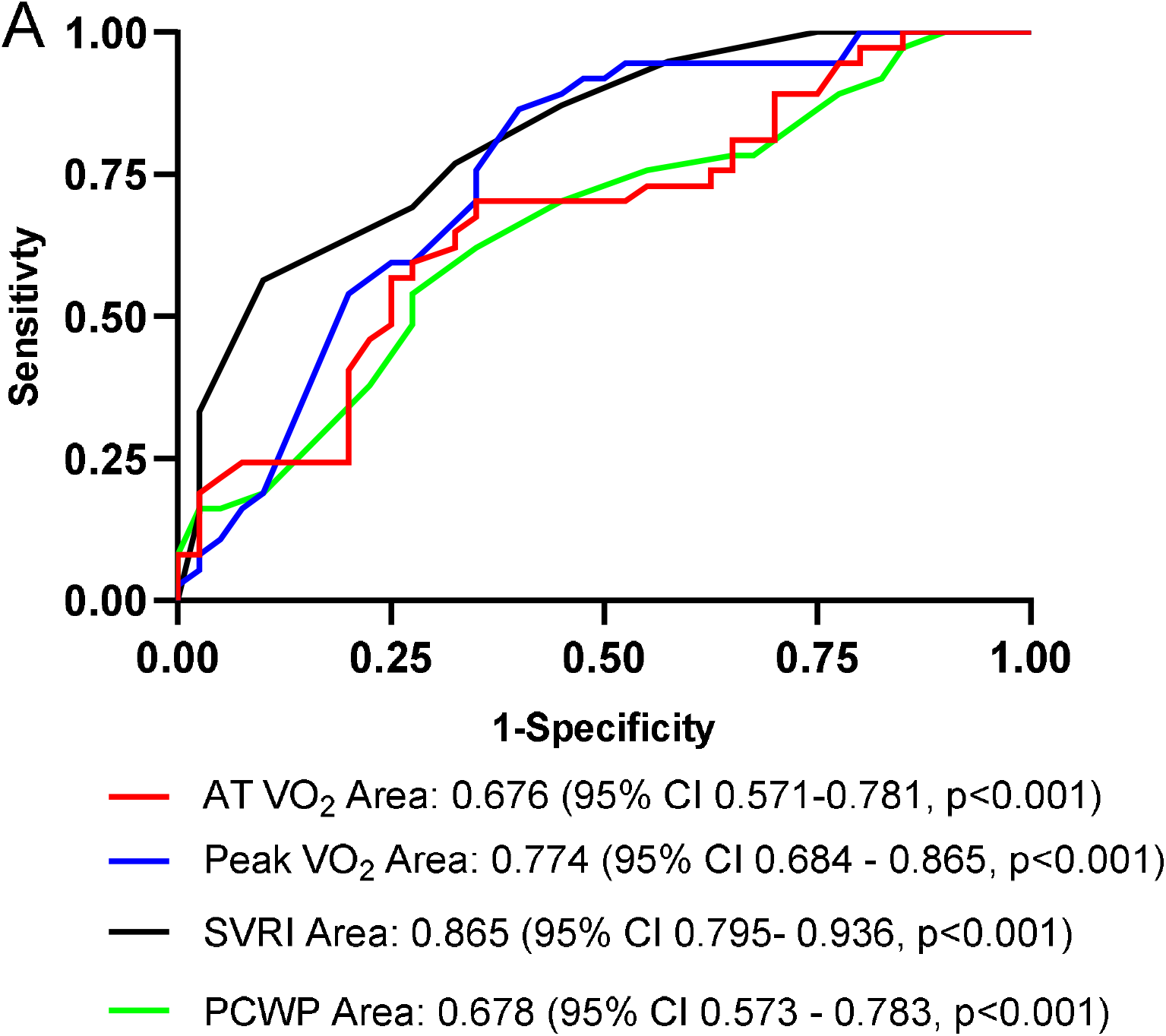
ROC curves for AT VO_2_, peak VO_2_, SVRI, and PCWP. The C-statistic was highest for SVRI compared to other single-predictors. AUC (C-statistic) and the respective 95% CI are listed. VO_2_, oxygen consumption; AT VO_2_, VO_2_ at anaerobic threshold; SVRI, stroke volume reserve index; PCWP, pulmonary capillary wedge pressure; AUC, area under the curve; CI, confidence interval.

Figure 3 shows cross-classification into four cohorts using SVRI and peak VO_2_. The top two cohorts with the best survival were defined by normal SVRI while the bottom two cohorts with the worst survival were defined by low SVRI.

**Figure 3.**
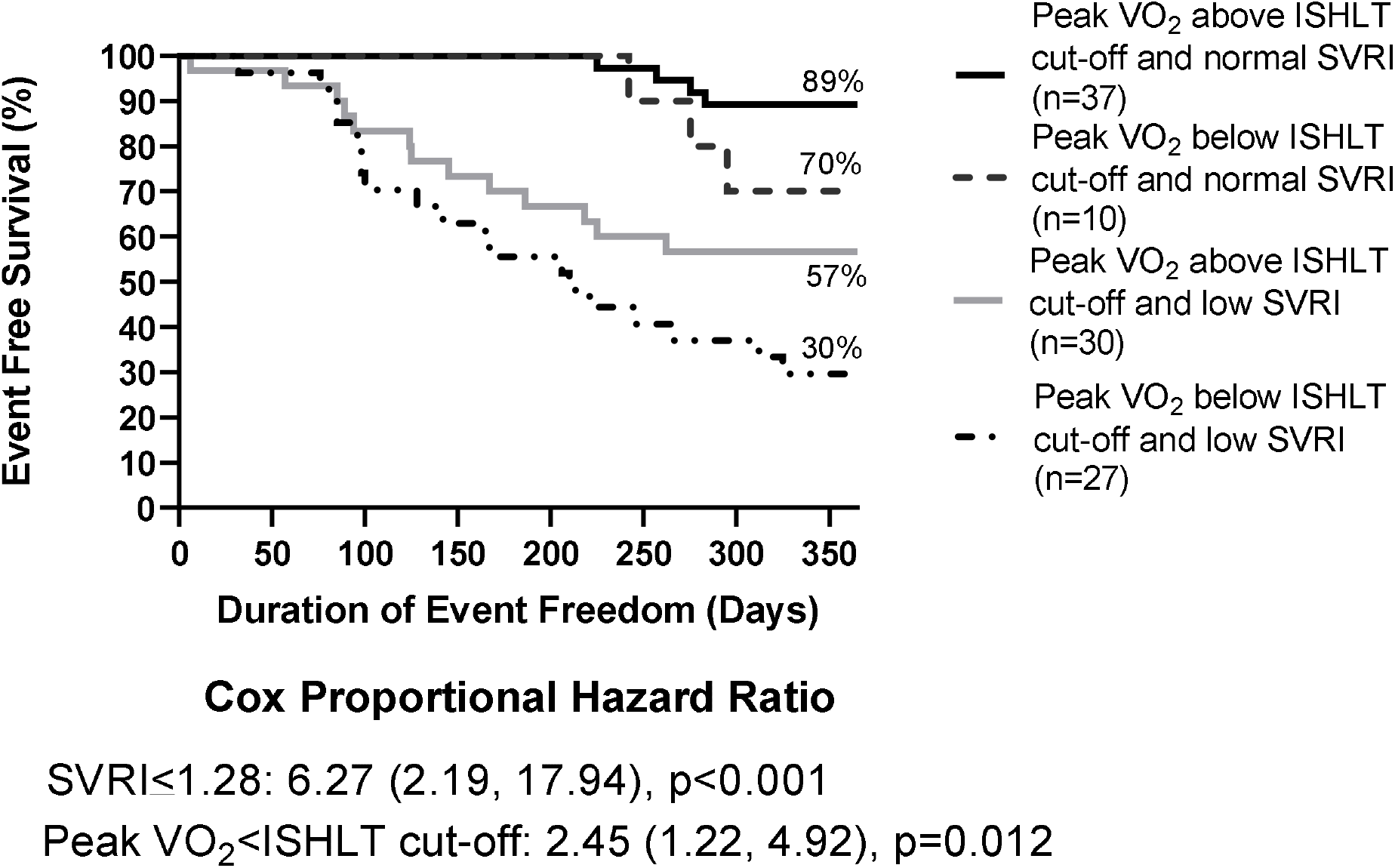
Kaplan-Meier survival curves showing the cross-classification of the patient population into four groups based on the peak VO_2_ below/above ISHLT cut-off and SVRI below/above 128%. Patients with normal SVRI showed the best event free survival regardless of their peak VO_2_. Within the HFrEF patients with a peak VO_2_ above the ISHLT cut-off, patients with an low SVRI showed a significantly worse 1-year event free survival compared to patients with normal SVRI. The separation of event free survival is again seen early at 100 days. SVRI≤128% and peak VO_2_ below the ISHLT cut-off were analyzed in a multi-predictor Cox proportional hazards model to derive the hazard ratios listed in the figure and their corresponding 95% confidence intervals—the reference group was SVRI>128% and peak VO_2_ above the ISHLT cut-off. VO_2_, oxygen consumption; ISHLT, International Society for Heart and Lung Transplantation; SVRI, stroke volume reserve index; HFrEF, heart-failure with reduced ejection fraction.

Patients with peak VO_2_ above the 2016 ISHLT cut-off and low SVRI demonstrated similar prognosis to HFrEF patients with peak VO_2_ below the ISHLT cut-off (57% vs 42%, p = 0.37; supplemental fig. S1). HFrEF patients with a peak VO_2_ below the ISHLT cut-off but normal SVRI showed similar prognosis to HFrEF patients with peak VO_2_ greater than the ISHLT cut-off (70% vs 74%, p = 0.95; supplemental fig. S2).

### SVRI as a Predictor Among Patients with Adequate Peak VO_2_

Among the entire group, Cox proportional hazards analysis disclosed that low SVRI (HR 6.27, 95% CI 2.19, 17.94, p<0.001) and peak VO_2_ below the ISHLT cut-off (HR 2.45, 95% CI 1.22, 4.92, p=0.012) were both independently predictive of the clinical outcome. Among only those patients with “adequate” peak VO_2_ by ISHLT guidelines, patients with low SVRI had significantly worse 1-year event free survival than those with high SVRI (57% vs 89%, p = 0.001) (Fig. 3).

### Single-Predictor and Multi-Predictor Logistic Regression of Candidate Predictors of Adverse Outcome

The continuous variables, higher NYHA functional class, lower SVRI, higher VE/VCO_2_ slope, lower AT VO_2_, lower peak VO_2_ pulse, lower HFSS score and decreased hemoglobin levels were found to be significantly associated with the primary outcome (Table 2). In addition, several hemodynamic parameters obtained from right heart catheterization, including higher mean RAP, higher MPA, higher PCWP, lower mixed venous oxygen saturation and decreased resting cardiac index, were associated with 1-year outcome. The categorical variable of the Kato model score four or greater significantly correlated with 1-year outcome, while the SHFM score predicting < 80% 1-year survival did not. Subsequently, SVRI, peak VO_2_, AT VO_2_, VE/VCO_2_ slope, peak oxygen pulse, mean RAP, MPA, PCWP, resting cardiac index, mixed venous oxygen saturation, hemoglobin level, and NYHA class were selected for inclusion in the multi-predictor model. Stepwise backward logistic regression was used to arrive at the final predictive model (table 3)—notably SVRI≤128%, peak VO_2_<12 to 14 ml/kg/min or ≤ 50% predicted as per the 2016 ISHLT heart-transplant listing guidelines^7^, and resting cardiac index≤2.0 were independently associated with clinical outcomes. The OR using the pre-defined cut-off of SVRI≤128% was highest at 6.7.

**Table 2.** Factors Associated With 1-Year Composite Events in Single-Predictor Logistic Regression

| Variables | OR (95% CI) | p-value |
| --- | --- | --- |
| Age (years) | 1.022 (0.976, 1.071) | 0.35 |
| NYHA class | 4.552 (1.713, 12.098) | 0.002 |
| SVRI | 0.0173 (0.002, 0.143) | <0.001 |
| Peak VO<sub>2</sub> (ml/kg/min) | 0.736 (0.627, 0.863) | <0.001 |
| AT VO<sub>2</sub> (ml/kg/min) | 0.801 (0.672, 0.954) | 0.013 |
| VE/VCO<sub>2</sub> slope | 1.116 (1.030, 1.188) | 0.006 |
| Peak oxygen pulse (ml/beat) | 0.781 (0.683, 0.920) | 0.003 |
| Mean RAP (mmHg) | 1.115 (1.023, 1.217) | 0.013 |
| Mean PA (mmHg) | 1.045 (1.003, 1.088) | 0.034 |
| PCWP (mmHg) | 1.066 (1.010, 1.125) | 0.020 |
| RVSWI (g·m<sup>2</sup>/beat) | 0.911 (0.796, 1.042) | 0.17 |
| PVR (Wood units) | 0.977 (0.893, 1.070) | 0.62 |
| TPG (mmHg) | 1.041 (0.938, 1.155) | 0.45 |
| Mixed venous oxygen saturation (%) | 0.890 (0.833, 0.948) | <0.001 |
| Cardiac index (liters/min/m<sup>2</sup>) | 0.207 (0.072, 0.594) | 0.003 |
| Hemoglobin (mmol/L) | 0.583 (0.384, 0.884) | 0.011 |
| Sodium (mmol/L) | 0.900 (0.794, 1.018) | 0.095 |
| Creatinine (μmol/L) | 1.004 (0.996, 1.012) | 0.317 |
| HFSS | 0.298 (0.155, 0.587) | 0.005 |
| SHFM* | 1.68 (0.572, 4.921) | 0.35 |
| Kato model score* | 8.438 (1.686, 42.223) | 0.009 |

NYHA, New York Heart Association; SVRI, stroke volume reserve index; VO<sub>2</sub>, oxygen consumption; AT VO<sub>2</sub>, VO<sub>2</sub> at anaerobic threshold; VE/VCO<sub>2</sub>, minute ventilation/carbon dioxide output; RAP, right atrial pressure; PA, pulmonary artery pressure; PCWP, pulmonary capillary wedge pressure; RVSWI, right ventricular stroke work index; PVR, pulmonary vascular resistance; TPG, transpulmonary pressure gradient; HFSS, Heart Failure Survival Score; SHFM, Seattle Heart Failure Model; CI, confidence interval; OR, odds ratio.
\* Indicates a categorical variable.

**Table 3.** Multi-predictor Logistic Regression of Factors Associated with 1-Year Events. Covariates were dichotomized using published or data-based cut-offs.

| Predictors | OR (95% CI) | *p*-value |
| --- | --- | --- |
| SVRI≤128% | 6.690 (2.08, 21.49) | 0.001 |
| Cardiac Index≤2.0 | 3.176 (1.26,10.92) | 0.017 |
| Peak VO<sub>2</sub> below 2016 ISHLT cut-off | 4.152 (1.35,12.75) | 0.013 |

SVRI, stroke volume reserve index; VO<sub>2</sub>, oxygen consumption; ISHLT, International Society for Heart and Lung Transplantation; CI, confidence interval; OR, odds ratio.

### Predictive Model and Risk Score Generation

We subsequently created a novel SVRI risk score in which SVRI≤128% is weighted 2 points, and peak VO_2_ below the ISHLT cut-off and resting cardiac index≤2.0 were both weighted 1 point. A receiver operator curve of the SVRI risk score disclosed a C-statistic of 0.886 (95% CI 0.819, 0.943) for association with 1-year events after CPET (Fig. 4). A cut-off of 3 points or greater yielded a sensitivity of 63.8%, a specificity of 80.6% and a predictive accuracy of 73.8%.

**Figure 4.**
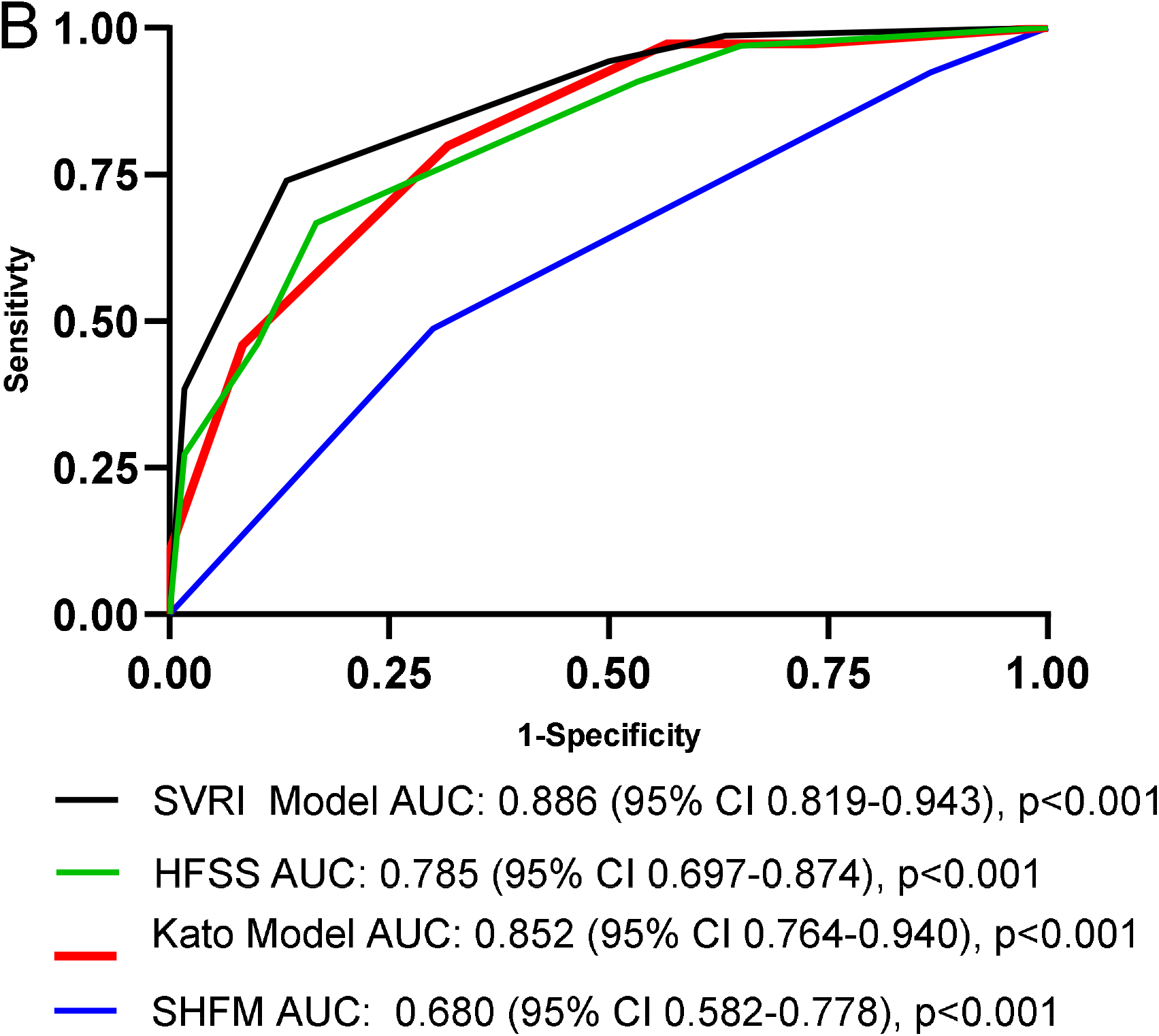
ROC curves for the SVRI risk score, HFSS, Kato model, and SHFM scores. The C-statistic was highest for the SVRI risk score compared to other risk score models. AUC (C-statistic) and the respective 95% CI are listed. ROC, receiver-operating-characteristic; SVRI, stroke volume reserve index; HFSS, Heart Failure Survival Score; SHFM, Seattle Heart Failure Model; AUC, area under the curve; CI, confidence interval.

## DISCUSSION

Our results disclosed that SVRI, a novel CPET parameter, independently predicted serious deterioration (death, heart-transplantation or urgent LVAD implantation) among HFrEF patients, with “excellent” to “outstanding” accuracy.^33^ SVRI provides prognostic information more accurate than currently recommended CPET parameters, such as peak VO_2_, AT VO_2_, and VE/VCO_2_ slope. Ambulatory HFrEF patients with low SVRI had a little over half the 1-year survival rate of those with normal SVRI. Notably, low SVRI also predicted a poor 1-year prognosis among HFrEF patients with otherwise “adequate” peak VO_2_. Based on our results, we created a simple risk-stratification model based in large part on the SVRI that reliably identified patients with a high risk for 1-year events in this cohort.

During the initial and intermediate phases of exercise, SV contributes to the increase in cardiac output, after which output primarily depends on heart rate.^17, 34^ Our method focuses on the critical exercise period between rest and AT and the consistent change in SV from rest to AT observed in normal patients.^16-19, 21^ We calculated SVRI during CPET performance without requiring changes to the standard protocol or patient instructions. SVRI was not shared with the advanced therapies selection committee and did not contribute to the committee’s decision for candidacy for heart-transplant or LVAD implantation.

Although peak VO_2_ during CPET in part reflects the capacity to increase cardiac output during exercise^35^, it is also influenced by noncardiac factors such muscle deconditioning, age, gender, race, obesity, anemia, lung function and use of (3-blockers.^36^ Perhaps for this reason, peak VO_2_ has had a limited ability to distinguish HFrEF patients with imminent risk of deterioration from those with better prognoses.^22, 37, 38^ For example, the C-statistic of peak VO_2_ for detecting deterioration among various groups of heart failure patients was only 0.62–0.69, below the discriminatory limit commonly considered acceptable.^33^ Other CPET parameters such as peak VO_2_ pulse, VE/VCO_2_ slope and AT VO_2_ are similarly limited in accuracy.^39^

SVRI has other important advantages. It is a noninvasive test of inotropic reserve. Since the measurements are completed once the patient reaches AT, it does not rely on maximal effort, which can vary particularly among HFrEF patients.^39^ In addition, SVRI performed well in our study without the requirement for age-, sex-, race-matched controls for determination of abnormal values.

### Risk-stratification in HFrEF Patients

Currently, there are no universal criteria for identifying HFrEF patients who should be evaluated for advanced heart-failure therapies.^4^ Consequently, risk scores such as the SHFM and HFSS are often used to identify HFrEF patients at high risk for 1-year events. For this study, we chose to evaluate the SVRI risk score with SHFM and HFSS as they are the most commonly recommended multivariable risk scores by the ISHLT, American College of Cardiology, and European Society of Cardiology.^7, 40, 41^ Additionally, we evaluated the Kato model score as this is a more recent risk score that uses peak VO_2_ and markers of left and right sided congestion to guide risk assessment.^31^ In our patient population, the HFSS and Kato model score showed predictive value while the SHFM did not. The SVRI risk score shows a trend to superior predictive accuracy compared to both HFSS and Kato model scores. Thus, in a patient population where multivariable risk scores historically have shown only moderate accuracy^42^, we demonstrate the incremental value of utilizing SVRI.

Notably, SVRI is valuable in identifying HFrEF patients who have “adequate” peak VO_2_ (by ISHLT criteria) but who are nevertheless at high risk of deterioration. Figure 5 demonstrates a useful schematic for cross-classification of HFrEF patients by SVRI and peak VO_2_. Additionally, HFrEF patients who were initially considered to be high risk by peak VO_2_, but who demonstrate normal inotropic reserve by SVRI have similar survival to patients with a peak VO_2_ greater than the ISHLT cut-off. We speculate that HFrEF patients with low peak VO_2_ but normal SVRI have noncardiac reasons that contribute to the low peak VO_2_, such as deconditioning and anemia, which may improve with interventions such as cardiac rehabilitation and improved nutritional status. Thus, SVRI can potentially identify patients who will subsequently show improvement in peak VO_2_, VE/VCO_2_ slope and AT VO_2_ with pharmacological and non-pharmacological approaches.^43-46^ More importantly, low peak VO_2_ with normal SVRI might identify patients who would not necessarily benefit from heart-transplantation or LVAD implantation because of their preserved inotropic reserve. Thus, cross-classification of HFrEF patients using both SVRI and peak VO_2_ may identify patients with “reversible” noncardiac factors (low peak VO_2_ with normal SVRI) while identifying patients with poor inotropic reserve who remain at high risk for 1-year events despite their seemingly adequate exercise tolerance (adequate peak VO_2_ with low SVRI). Our SVRI risk score includes SVRI, peak VO_2_ and resting cardiac index and thus, we believe shows improved accuracy because it separately accounts for both cardiac and noncardiac factors that would lead to death, heart-transplantation and urgent LVAD implantation.

**Figure 5.**
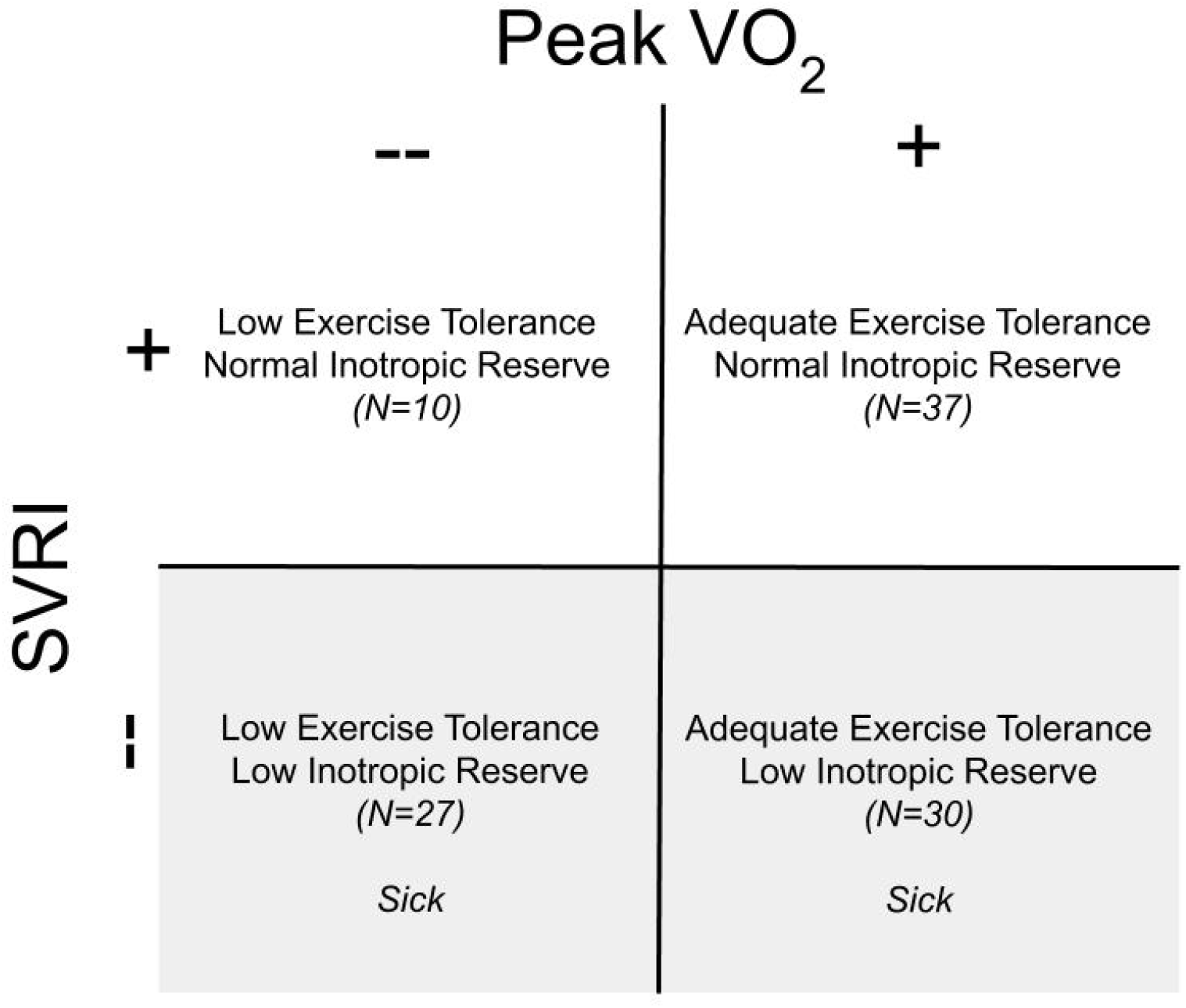
Schematic using cross-classification of heart-failure with reduced ejection fraction patients by SVRI below/above 128% threshold and peak VO_2_ below/above 2016 ISHLT cut-off. SVRI, stroke volume reserve index; VO_2_, oxygen consumption; ISHLT, International Society for Heart and Lung Transplantation.

### Methodologic Considerations and Future Directions

Our study contains several limitations. First, this is a single center retrospective cohort study and thus our observations need to be validated in other centers with larger patient samples to confirm our results. Our patient population was predominantly older males with coronary artery disease but not dissimilar to other studies evaluating HFrEF patients.^2, 11, 31^ Second, we did not confirm our estimates of SV with noninvasive nor invasive determinations of CO. Confirmation with direct measures of CO would demonstrate whether inappropriate increase in CO is the underlying reason for why SVRI is able to identify patients at high risk for 1-year events. However, our study uses CPET protocols commonly used in other cardiac centers and thus demonstrate the potential generalizability of our observations. Third, we did not include numerous other multivariable risk scores for comparison as they have shown similar results with moderate accuracy and a C-statistic consistently in the 0.7 to 0.8 range.^42, 47^ Our goal was to compare the SVRI risk score with currently recommended multivariable risk scores, particularly those that include the use of peak VO_2_ in their risk calculation. Finally, this study was conducted using treadmill exercise only (preferred modality in the United States^24^) and thus it is unknown whether the same conclusions can be made in CPETs using a stationary cycle ergometer.

The “SVRI risk score” was generated internally from our own data, so would need to be externally validated in another population of patients with HFrEF. We speculate that the SVRI and SVRI risk score would provide useful prognostic information in other heart-failure patient populations such as heart-failure with preserved ejection fraction and congenital heart disease patients, but further studies would be necessary to confirm its performance. Additionally, it would be useful to evaluate whether SVRI can accurately identify HFrEF patients who can significantly increase their peak VO_2_ after cardiac rehabilitation. Finally, this study was conducted during the transition to the heart allocation change by the United Network of Organ Sharing that took place in the United States in November 2018. Future studies will have to take this into consideration as we would expect it to change the time to heart-transplantation in ambulatory HFrEF patients.

## CONCLUSION

SVRI is a novel CPET parameter that accurately predicts 1-year prognosis in HFrEF patients. Our SVRI risk score using cut-off values consisting of SVRI≤128%, peak VO_2_ below the ISHLT cut-off, and resting cardiac index≤2 suggests improved prognostic accuracy compared to currently recommended multivariable risk scores. Therefore, SVRI should be considered when evaluating HFrEF patients for advanced therapies.

## Data Availability

The data referred to in the manuscript is available and can be obtained by emailing

## ACKNOWLEDGMENTS

We would like to thank Dr. Barry Greenberg for his language editing and proofreading of the manuscript.

---

**Figure S1.**
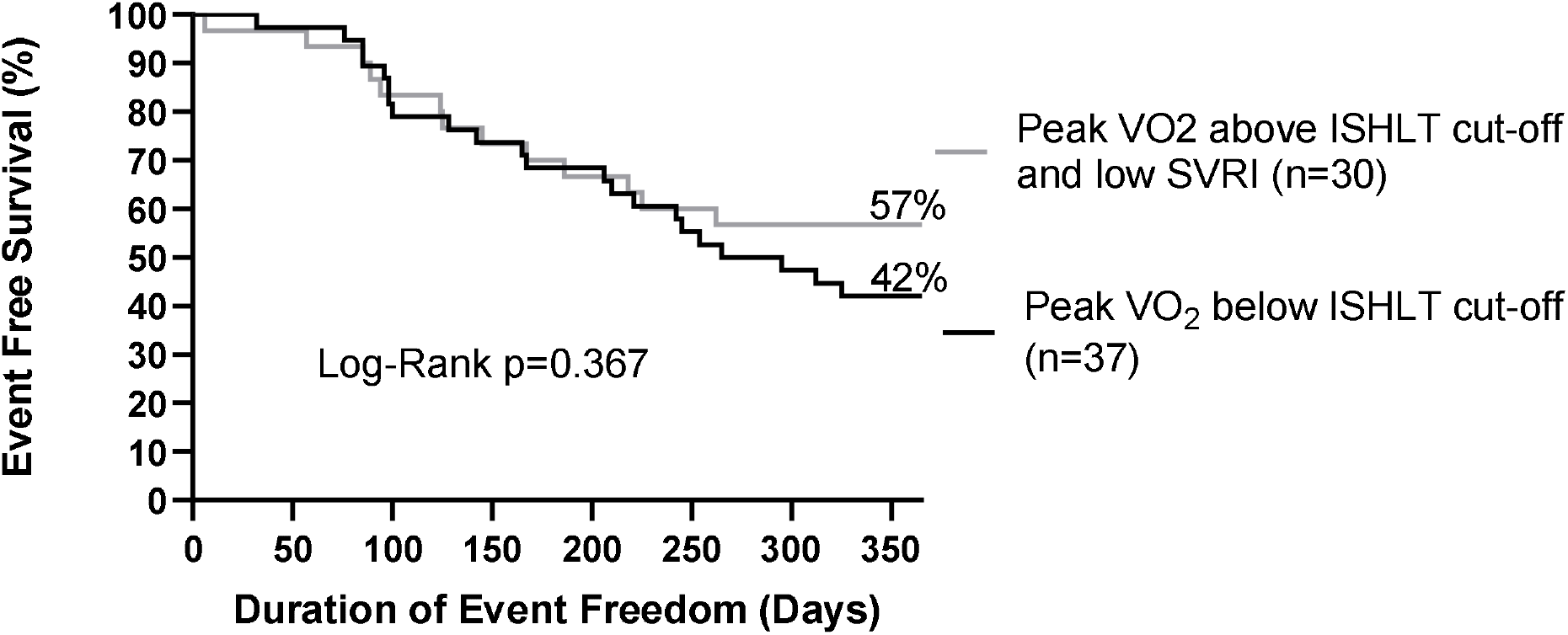
Kaplan-Meier survival curves of heart-failure with reduced ejection fraction patients with peak VO_2_ below the ISHLT cut-off (solid line) in comparison to patients with peak VO_2_ above the ISHLT cut-off and with low SVRI (gray line). No significant differences were present in the event free survival between the two groups. VO_2_, oxygen consumption; SVRI, stroke volume reserve index; ISHLT, International Society for Heart and Lung Transplantation.

**Figure S2.**
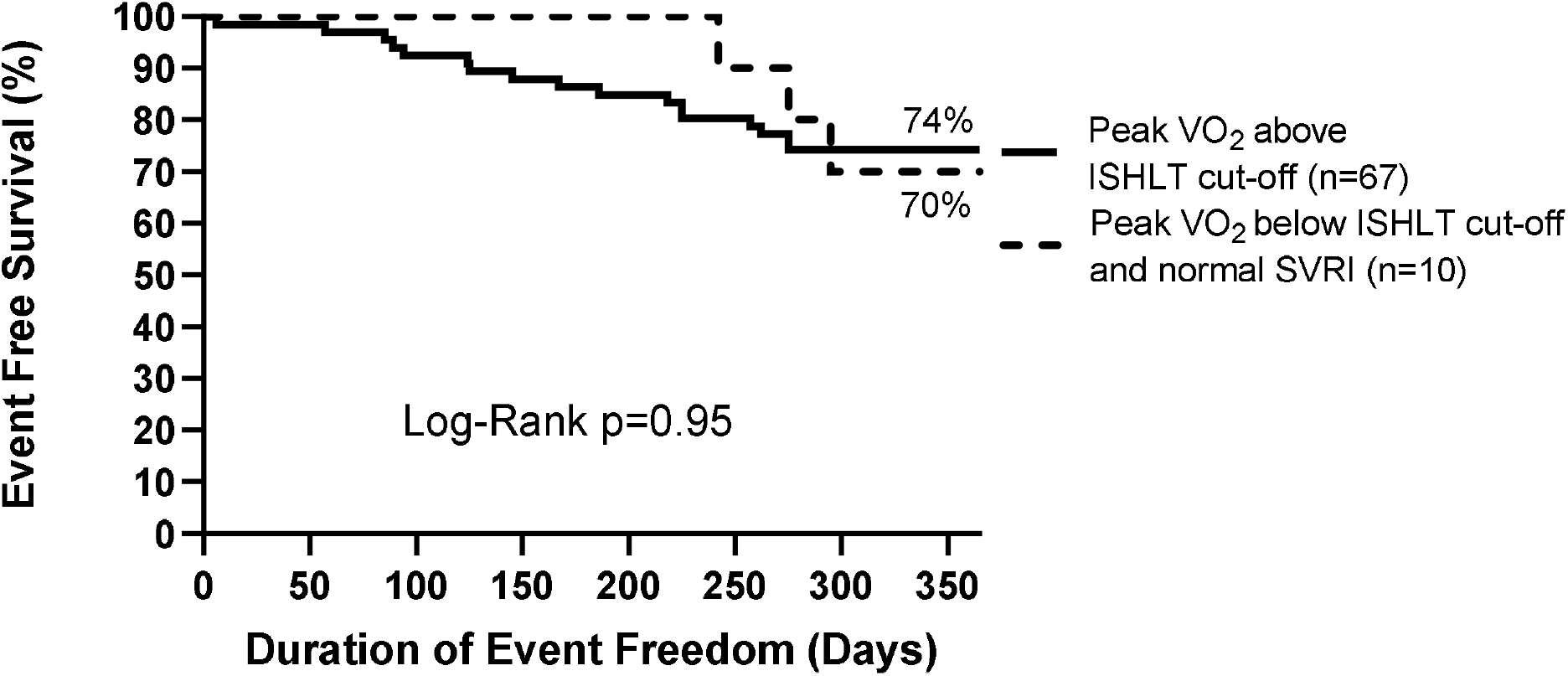
Kaplan-Meier survival curves of heart-failure with reduced ejection fraction patients with peak VO_2_ greater than the ISHLT cut-off (solid black line) in comparison to patients with peak VO_2_ below the ISHLT cut-off and with normal SVRI (dotted black line). No significant differences were present in the event free survival between the two groups. VO_2_, oxygen consumption; SVRI, stroke volume reserve index; ISHLT, International Society for Heart and Lung Transplantation.

